# Microsatellite Instability Detection in Clinical Cancer Samples: A Multiplex qPCR Approach without Matching Normal Samples

**DOI:** 10.1101/2023.11.07.23298217

**Authors:** Wei Chen, Yan Helen Yan, Blake Young, Alessandro Pinto, Qi Jiang, Nanjia Song, Weijie Yao, David Yu Zhang, Jinny Xuemeng Zhang

## Abstract

**Background:** Microsatellite instability (MSI) indicates DNA mismatch repair deficiency in cancers like colorectal cancer. The current gold standard technique, PCR/capillary electrophoresis (CE), requires matching normal samples and specialized instrumentation. We developed VarTrace, a rapid and low-cost quantitative PCR (qPCR) assay, to evaluate MSI using solely the tumor sample DNA, obviating the requirement for matching normal samples.

**Methods:** 101 formalin-fixed paraffin-embedded (FFPE) tumor samples were tested using VarTrace and compared to the Promega OncoMate assay utilizing PCR-CE. Tumor percentage limit of detection was evaluated on contrived samples derived from clinical MSI-H samples. Analytical sensitivity, specificity, limit of detection and input requirements were assessed using synthetic commercial reference standards.

**Results:** VarTrace demonstrated 100% test success rate, 100% sensitivity and 98% specificity compared to OncoMate across 101 clinical FFPE samples. It detected MSI-H with 97% accuracy down to 10% tumor percentage. Analytical studies using synthetic samples showed a limit of detection of 5% variant allele frequency and a limit of input of 0.5 ng.

**Conclusions:** This study validates VarTrace as a swift, accurate and economical assay for MSI detection in samples with low tumor percentages without the need for matching normal DNA. VarTrace’s capacity for highly sensitive MSI analysis holds potential for enhancing the efficiency of clinical workflows and broadening the availability of this crucial test.

## Introduction

Microsatellite instability (MSI) is characterized by length changes in repetitive DNA sequences called microsatellites and is often caused by deficiencies in DNA mismatch repair (dMMR) (1, 2). MSI is an important biomarker in several cancers (3, 4), most notably colorectal cancer (1, 5) where approximately 15% of patients are classified as MSI-H (6). MSI can indicate dMMR tumors and is also an important screening factor for Lynch syndrome (6–8), an inherited disorder caused by germline mutations in MMR genes that confers high lifetime cancer risks. Patients with high MSI (MSI-H) tumors have favorable prognosis and derive great benefit from immunotherapy compared to aggressive chemotherapy(9–17).

Current standard methods for detecting MSI and dMMR in colorectal cancer include immunohistochemistry (IHC), PCR amplification of microsatellite markers followed by capillary electrophoresis (PCR-CE) (18, 19), or next-generation sequencing (NGS) (20–28) as recommended by the National Comprehensive Cancer Network. IHC can reflect MSI status through identifying loss of MMR protein expression but may miss some MSI tumors with intact protein expression (29, 30). NGS and PCR-CE directly classify tumors as MSI-H or microsatellite stable (MSS) but often require comparison to matched normal tissue (18–26). While NGS can accurately assess MSI and MMR gene status from minute inputs (25), it requires specialized instrumentation. PCR-CE is considered the gold standard for MSI detection due to its high efficiency and sensitivity in detecting repeat markers. The Bethesda Guidelines recommend a panel of five well-characterized homopolymer markers for MSI testing by PCR-CE: BAT-25, BAT-26, NR-21, NR-24, and MONO-27(31). However, neither IHC nor PCR-CE alone ensures accurate diagnosis (18, 29, 30, 32, 33). Using both methods simultaneously increases sample requirements and screening costs substantially. Moreover, the multi-step PCR-CE process still demands advanced instrumentation, with only marginal cost savings over NGS. Alternative methods such as droplet digital PCR (32) and targeted next-generation sequencing panels (25–28) have also emerged for accurate MSI screening, but require further validation.

To reduce costs and sample input while maintaining sensitivity for MSI detection, we developed a rapid qPCR assay using specially designed primers and blockers to preferentially amplify MSI alleles over wildtype sequences. Our assay requires only 0.5 ng DNA input and can directly detect MSI in DNA samples extracted from just 10% tumor content without preamplification. Combined with automated data analysis, MSI status can be determined within 8 hours of receiving a sample, much faster than existing PCR-CE protocols. This qPCR approach could potentially be applied to both tumor tissue and cell-free DNA pending clinical validation. By enabling rapid, robust MSI detection from minute DNA quantities, this method could greatly expand test accessibility in labs with qPCR instrument and facilitate Lynch syndrome screening and precision medicine in colorectal cancer management.

## Material and Methods

### Samples

Formalin-fixed paraffin-embedded (FFPE) samples were obtained from several sources. 12 samples were purchased from BioChain Institute, of which 7 were confirmed to have MSI status by IHC and 5 were confirmed by Promega OncoMate MSI Dx assay. 41 samples with a diagnosis of adenocarcinoma of the7colon/rectum were purchased from OriGene, but MSI status was not determined. 48 samples with MSI status confirmed by PCR were purchased from Discovery Life Sciences. All clinical samples information is collected from vendors and summarized in Table 1 and Supplemental Table S1-3.

**Table1.** Sample information.

|  |  | **TOTAL** | **MSI-H<sup>1</sup>** | **MSI-L/**<br>**MSS<sup>1</sup>** | **NA** |
| --- | --- | --- | --- | --- | --- |
| **VARIABLES** |  | 101 | 45 | 50 | 6 |
| **SEX** | Female | 46 | 20 | 23 | 3 |
|  | Male | 41 | 12 | 26 | 3 |
|  | Unknown | 14 | 13 | 1 | 0 |
| **AGE** | >=50 | 59 | 19 | 37 | 3 |
|  | <50 | 12 | 3 | 8 | 1 |
|  | Unknown | 30 | 23 | 5 | 2 |
| **STAGE** | I | 12 | 9 | 3 | 0 |
|  | II | 25 | 18 | 7 | 0 |
|  | III | 31 | 13 | 14 | 4 |
|  | IV | 29 | 2 | 25 | 2 |
|  | Unknown | 4 | 3 | 1 | 0 |
| **TUMOR**<br>**TISSUE TYPE** | Primary | 73 | 44 | 25 | 4 |
|  | Metastatic | 28 | 1 | 25 | 2 |
| **TUMOR**<br>**LOCATION** | Colon | 61 | 40 | 18 | 3 |
|  | Rectum | 5 | 1 | 3 | 1 |
|  | Other (cecum, small intestine, stomach, liver) | 32 | 1 | 29 | 2 |
|  | Unknown | 3 | 3 | 0 | 0 |
| **TUMOR**<br>**ORIGIN** | Colon | 87 | 41 | 42 | 4 |
|  | Rectum | 7 | 1 | 4 | 2 |
|  | Other (cecum, small intestine, stomach) | 4 | 0 | 4 | 0 |
|  | Unknown | 3 | 3 | 0 | 0 |

1. FFPE sample information is categorized by MSI status detected using Promega OncoMate MSI Analysis, as this is the current 'gold standard'. FFPE samples are all commercially purchased from vendors, focusing on with samples with known MSI status from BioChain and Discovery, and samples from colorectal or other related cancer from OriGene.

Seraseq® MSI Reference Panel Mix AF5% (Seracare, catalog number: 0710-1675) was used as Commercial reference standard for sensitivity, specificity and limit of input study. In addition, VarTrace Positive and Negative Controls was also included as a comparison in for sensitivity, specificity and limit of input (LoI) study. VarTrace Negative Control is background human genomic DNA GM25485, which is known to be microsatellite stable (MSS). VarTrace Positive Control is a mixture of GM25485 and 5 synthetic MSI-H DNA fragments, and variant for each marker was quantified as ∼5% variant allele frequency (VAF) using qPCR individually. DNA fragments used in Positive Control are plasmid synthesized and purified in GenScript. Additional WT and variants templates used in limit of blank (LoB) and limit of detection (LoD) study respectively are synthetic gBlock fragments ordered from IDT, sequences are listed in Supplemental Table S4.

### NuProbe VarTrace MSI assay

The VarTrace MSI Kit comprises 2 tubes of 4x oligo mixes that can detect totally 5 markers, a Positive Control and a Negative Control. The background DNA used in both Positive and Negative Controls is cell line GM25485 (BioChain), which is known to be MSS. TaqPath ProAmp Multiplex Master Mix (Applied Biosystems, Catalog number: A30868) was used to conduct all VarTrace experiment. All VarTrace experiment was prepared strictly following the user manual (Supplemental Methods) and perform qPCR readout using Applied Biosystems 3500 Fast Dx (Table 2).

**Table2.** Comparison of Operating Characteristics and Specification between Promega Oncomate and NuProbe VarTrace.

|  | Promega OncoMate | NuProbe VarTrace |
| --- | --- | --- |
| Normal Sample Required | Yes | No |
| Instrument Setting<sup>1</sup> | ABI 7500 Fast (qPCR) **AND**<br>ABI 3500 Dx (Sanger) | ABI 7500 Fast (qPCR) |
| Sample throughput<sup>2</sup> | 12 | 46 |
| Time to result | 2-day result | Same-day result |
| Tumor Perc of FFPE Sample | >=30% | >=10% |
| Input requirement | 1 ng | 0.5 ~ 10 ng |
| Success Rate | 94% | 100% |

1. ABI 7500 Fast (qPCR) is short for Applied Biosystems™ 7500 Fast Dx Real-Time PCR; ABI 3500 Dx (Sanger) is short for Applied Biosystems™ 3500 Dx Genetic Analyzer.
2. Sample throughput is calculated based on instrument specification.
ABI 3500 Dx (Sanger) throughput is 24 capillary per run, including matched sample. So, results are available for 12 patient per run per instrument
ABI 7500 Fast (qPCR) throughput is 96 wells per run, including 2 tubes per sample. Two Positive Control in plate will reduce the throughput. So, results are available for up to 46 patient per run per instrument.

### MSI Testing Using PCR-CE Method

Promega Oncomate MSI Dx is an assay cleared by FDA to determine MSI status, and well known as the “gold standard’. All FFPE samples underwent MSI testing using the Promega OncoMate MSI Dx Analysis assay per the manufacturer’s protocol. Testing was performed by a third-party CLIA-certified lab, which carried out DNA extraction, quantitation, and Promega assay on the samples. Redundant DNA was returned to NuProbe and stored at 4°C for 2 years prior to additional test.

### DNA Extraction and Quantification

For OriGene and BioChain samples, DNA was extracted from one slide per sample and no tumor preselection using the GeneRead FFPE Kit (Qiagen, Germany, Catalog number: 180134) per manufacture protocol with a slight modification (details in Supplemental Methods).

Considering only one slide per sample was obtained from Discovery, DNA extraction was performed in the third-party CLIA lab for Promega OncoMate assay test. Redundant DNA was returned and stored at 4°C. Prior to VarTrace Assay test, part of Discovery DNA samples was purified and concentrated for a second time due to magnetic bead residue.

All samples are quantified for DNA concentration prior to VarTrace tests using Invitrogen Qubit 4 Fluorometer (ThermoFisher Scientific, Catalog number: Q33238) and Qubit™ dsDNA Quantification Assay, High Sensitivity (ThermoFisher Scientific, Catalog number: Q32851).

### Data Analysis

The QuantStudio 3/5 Real-Time PCR Design & Analysis 2 software is used to extract Cq values under default settings. In the analysis pipeline (Supplemental Method and Fig. S1), samples are labeled as failed if the Cq value of *GAPDH* is larger than 37 or no amplification occurred. For each sample and control, Cq Value of *GAPDH* is subtracted from Cq value of each marker obtain 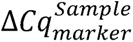. A marker is considered positive if 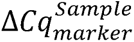 is smaller than 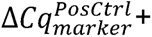 1. Samples with more than 2 positive markers were classified as MSI-H, those with 1 positive marker as MSI-L, and those with 0 positive markers as MSS. Both MSI-L and MSS are considered as Negative in final report, and only MSI-H is considered as Positive.

## Results

### Blocker displacement amplification enables qPCR detection of MSI at Low Variant Allele Frequencies

NuProbe developed VarTrace qPCR assay to detect MSI using a previous published blocker displacement amplification (BDA) technology without matching normal sample. BDA can manipulate reaction thermodynamics by introducing a rationally designed blocker oligonucleotide that perfectly binds to wildtype (WT) targets and shares a same binding region with the primer (Fig. 1A). Blocker has a stronger binding affinity towards WT templates comparing to primer, resulting in the inhibition of primer binding and extension. On the other hand, bulge formed between variant template and blocker can weaken the blocker binding stability and better amplification. Therefore, BDA can detect low frequency variants by preferably amplify variants over WT templates and generating fluorescence signals in qPCR setting.

**Fig. 1.**
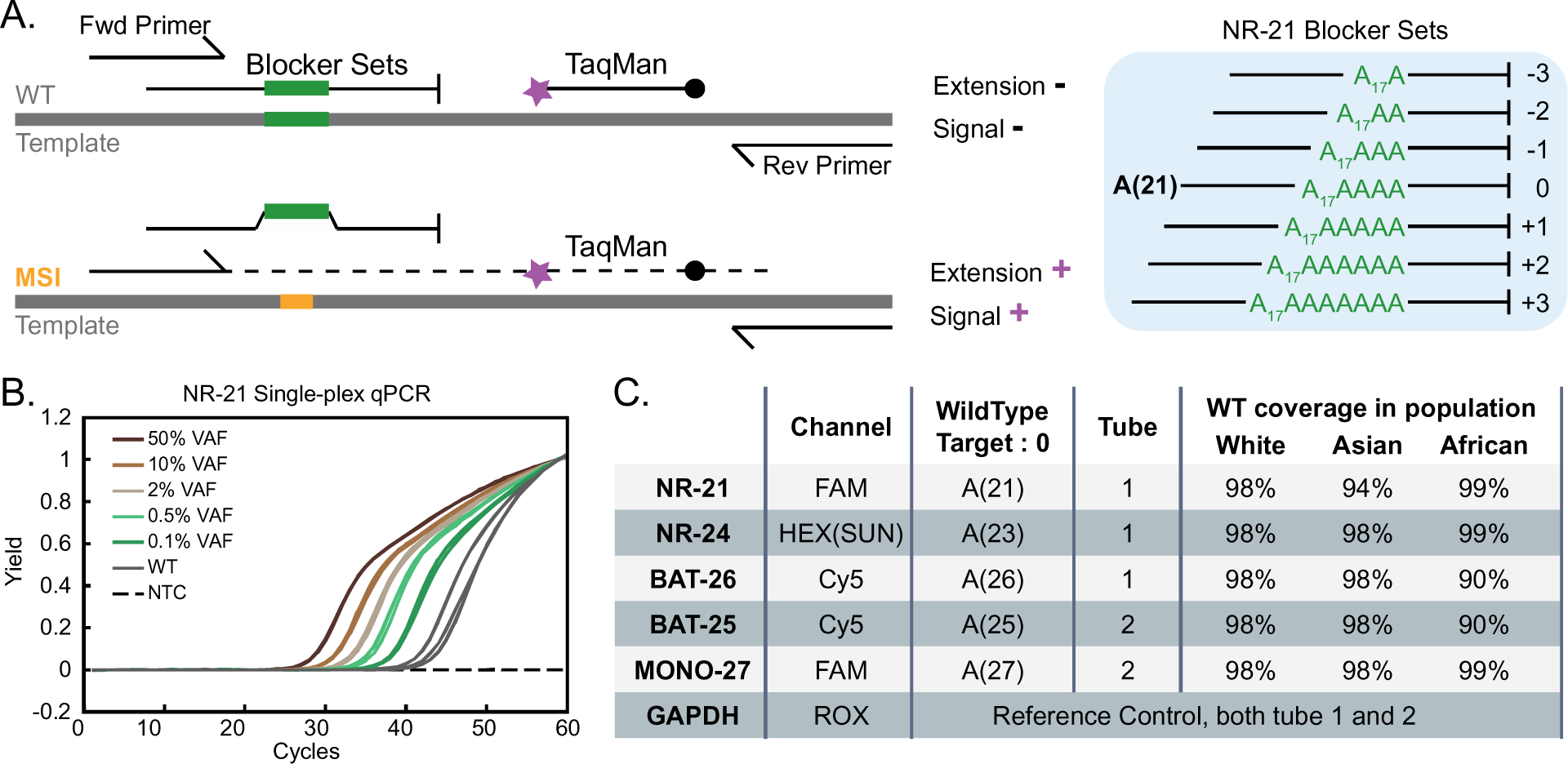
Detecting microsatellite instable variants using qPCR with BDA. (A) Single plex design diagram. Blocker sets (green) overlapping the forward primer binding site are designed to perfectly match common wildtype (WT) sequences: WT (0, ±1, ±2, ±3). This allows the blockers to outcompete forward primer binding to WT sequences. WT (0) represents the reference sequence with the highest population frequency, and WT (+1) represents sequences with a 1 nt addition in the mononucleotide repeat region relative to WT (0). MSI variants (red) create a mismatch that enables forward primer binding and PCR amplification. (B) qPCR detection of NR-21 variants in synthetic MSI-H samples with different variant allele frequencies (VAFs). WT (0) and no-template controls are shown in solid gray and dashed black, respectively. All variants and WT curves are tested and showed in triplicate, NTC is a single test. (C) Overview of the VarTrace assay composition and coverage. No blocker for *GAPDH*. Frequencies of genotypes covered by WTs (−3 to +3) were determined from population sequence data reported in previous publication (18).

The feasibility of single-plex qPCR with BDA in detecting microsatellite instability was first demonstrated by amplifying NR-21 unstable variants spiked into microsatellite stable (MSS) background DNA at variant allele frequencies (VAFs) down to 0.1% (Fig. 1B). There was significant separation of over 4 Cq cycles between the wildtype and 0.1% VAF samples, while no template controls showed no amplification. All 5 markers were tested individually with samples with a range of VAFs, and calibration curves for all 5 markers from 0.1% to 50% VAF showed high coefficients of determination (R^2) ranging from 0.93 to 0.99 (Supplemental Fig. S2), indicating both accurate detection and quantitation of MSI samples by qPCR using BDA.

Based on sensitive detection on each marker in single plex validation, VarTrace MSI assay comprises a set of seven blockers to target each one of 5 MSI markers from Bethesda guidelines: NR-21, NR-24, BAT-25, BAT-26 and MONO-27 (Fig. 1C). Primers targeting *GAPDH* are included as a reference control that reflects sample input and quality. *GAPDH* Cq value is utilized in the analysis algorithm. One set of blockers correspond to sequences of WT 0, ±1, ±2, and ±3 bp in order to minimize stutter products, short tandem repeats introduced by polymerase slippage. WTs (0) are selected to maximize coverage of natural variation in wild-type microsatellite lengths, enabling MSI detection without requiring a matched normal sample.

### Detection of MSI in FFPE Tumor Samples using the VarTrace Assay

A total of 101 FFPE tumor samples obtained from commercial sources were tested using the VarTrace assay, of which 60 had known MSI status from vendor. Successful DNA extraction, quantitation, and qPCR testing was achieved in the first round for all 53 samples extracted in-house. The remaining 48 samples from Discovery was extracted at an external provider, redundant DNA were sent back and stored at 4c for 2 years. Rehydration, centrifugation and magnetic separation were introduced to samples that have little liquid or with bead residues. All 48 samples were successfully tested after necessary procedure and DNA quantitation. No failed or invalid results occurred during screening of the 101 samples, and no retested needed.

The 101 samples encompassed a wide range of tumor percentages from 10% to 90% (median 40%). The VarTrace assay detected MSI-H in 47 samples, with tumor percentages ranging from 15% to 80% (median 45%) (Fig. 2A). Most MSI-H samples (40/47) were positive for ≥4 markers, while the remaining 7 samples with <50% tumor content were positive for only 2-3 markers (Fig. 2C). The MONO-27 marker showed no amplification in MSI-negative samples, while BAT-25 showed positive signals in all MSI-positive samples, indicating potential for optimization of marker specificity.

**Fig. 2.**
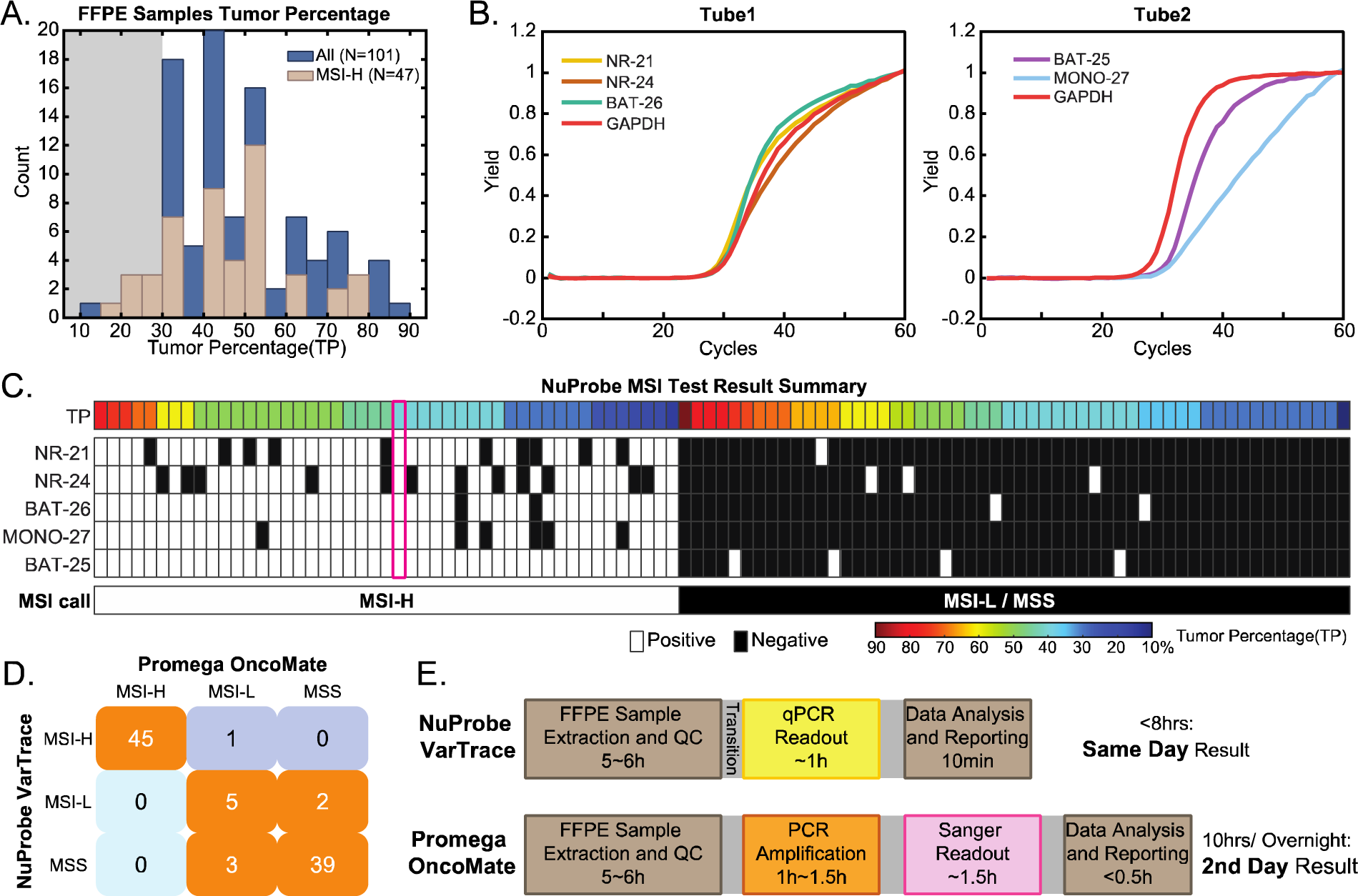
MSI Results of 101 FFPE tumor samples. Details on sample information are provided in the Methods and Table 1. (A) Tumor percentage distribution across all samples and MSI-H subset. Grey shading indicates samples below the 30% tumor percentage, which is below the Promega OncoMate specification for input samples. (B) Representative qPCR curves from an MSI-H sample (C504247) showing positive detection of all 5 markers, tube1 can detect 3 markers and the rest 2 markers are detected in tube2. *GAPDH* is present in both tubes and used as reference control. (C) Summary heatmap of MSI results from VarTrace for the 5 markers and overall MSI status. Samples are grouped by Positive (MSI-H) and Negative (MSI-L/MSS) status and sorted by decreasing tumor percentage within each group. White indicates positive detection or MSI-H status; black indicates no detection or MSI-L/MSS status. Sample C504247 in (B) is highlighted in magenta. (D) Concordance of MSI status between the 95 samples successfully tested by both VarTrace and OncoMate methods. 6 samples failed because of loss of marker signal in OncoMate tests. Diagnostic sensitivity (95% CI) is 100% (92%∼100%), and diagnostic specificity (95% CI) is 98% (89%∼100%). (E) Timeline comparison of workflow steps between VarTrace and OncoMate assay. See Table 2 for additional assay specification comparisons.

### Comparison of VarTrace Assay to Promega OncoMate MSI Analysis

The MSI status of the 101 FFPE samples was also tested using the Promega OncoMate assay at a third-party CLIA lab (Supplemental Table S5 and Supplemental Fig. S3-10). However, 6 samples showed invalid results due to marker readout failure, yielding a sampe test success rate of 94%. Of the remaining 95 samples, the VarTrace and OncoMate assays demonstrated significant concordance, with only 1 sample scored as MSI-H by VarTrace but MSI-L by OncoMate. The one discrepant sample was confirmed as MSI-H by the vendor report, suggesting OncoMate likely missed low-frequency MSI signals due to limitations in its pre-amplification efficiency and analysis algorithm (Supplemental Fig. S11). Considering both MSI-L and MSS as negative and MSI-H as Positive, the VarTrace assay demonstrated 99% concordance, 100% (95% CI 92%∼100%) sensitivity and 98% (95% CI 89%∼100%) specificity compared to OncoMate (Fig. 2D). Comparison of individual marker results between the VarTrace and Oncomate assays was also performed. The VarTrace assay demonstrated a specificity above 95% for 3 markers (NR-21, BAT-26, MONO-27) and a sensitivity and concordance above 95% for 3 markers (BAT-25, BAT-26, MONO-27).

### Comparison of VarTrace and OncoMate Assays to Vendor Validation

Of the 101 samples, 60 had MSI status validated by the commercial vendor, with 44 determined to be MSI-H. Compared to the vendor results, VarTrace concordantly classified 59/60 samples, with one sample scored as MSS by VarTrace but MSI-H by the vendor (Supplemental Table S5 and S6). This yielded a sensitivity of 98% (95% CI 88%∼100%) and specificity of 100% (95% CI 79%∼100%) for VarTrace.

Similarly, the OncoMate assay has 2 samples scored negative when vendor claimed to be MSI-H. After excluding 3 invalid samples from the validation dataset, the OncoMate assay demonstrated 95% sensitivity (95% CI 84%∼99%) and 100% specificity (95% CI 77%∼100%) versus vendor validation (Supplemental Table S6). Therefore, the VarTrace assay showed slightly better concordance with vendor results compared to OncoMate.

### Determination of Tumor Percentage Limit of Detection

Serial dilutions using wildtype DNA to make contrived samples at different tumor percentage were performed on 41 MSI-H samples (Fig. 3A and Supplemental Fig. S12-32), with each sample tested once at each dilution due to limited sample amount. The limit of detection of one sample is defined as the smallest tumor percentage that has positive result, and all tests with higher tumor percentage also report as positive. When evaluating limit of detection (LoD) of each individual sample, a tumor percentage LoD below 5% was achieved in 35/41 samples, with only 1 sample showing LoD above 10% (Fig. 3B and Supplemental Table S7). This sample originally carries a tumor percentage of 30% and tested positive in 3 markers, which may have sensitive marker performance and lose marker signal during serial dilution. All samples with 5 markers positive in undiluted DNA tests have tumor percentage LoD no bigger than 5%. Similarly, all samples with 4 markers positive at original testing showed tumor percentage LoD no bigger than 10%.

**Fig. 3.**
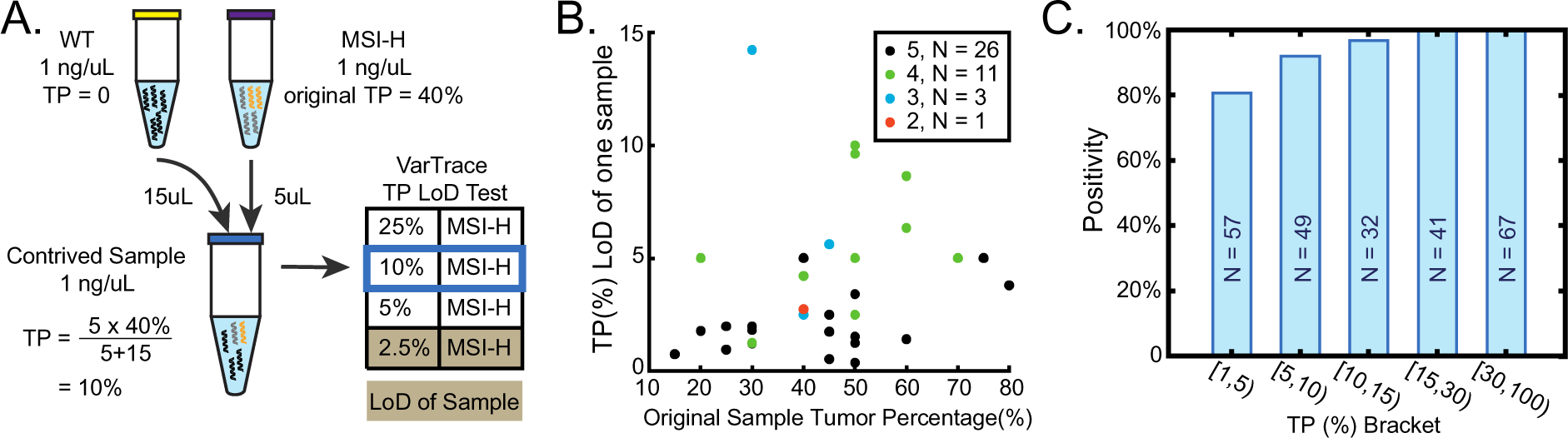
Tumor Percentage (TP) Limit of Detection (LoD). (A) Contrived sample dilution demonstration. Wildtype genomic DNA from cell line GM25485 was used as the MSS background (yellow tube) for all contrived sample dilutions. DNA extracted from an FFPE slide (C504247, 40% tumor percentage) was quantified by *GAPDH* in the original VarTrace test (Fig. 2B) and used as the starting MSI-H material (purple tube). The WT gDNA and FFPE DNA were mixed to create contrived samples at desired tumor percentage (blue tube). Serial dilutions were made to generate LoD datasets for this sample, with each dilution tested once. LoD was defined as the lowest tumor percentage at which all samples with ≥ LoD tumor percentage scored as MSI-H. (B) TP LoD determined by diluting 41 MSI-H samples with WT sample. Dot colors indicate number of positive markers detected from the undiluted sample. (C) Positivity of VarTrace assay across samples grouped into tumor percentage brackets, with number of samples per bracket showed in bar graph.

A total of 246 tests across varying tumor percentages were included in the analysis to evaluate the accuracy of the VarTrace assay at different tumor content levels, of which 41 tests using original undiluted sample are included. All results were considered equally, regardless of differences in starting material used to create the contrived diluted specimens. The positivity rate was calculated at different tumor percentage ranges to assess the performance of the VarTrace assay as tumor content decreased. VarTrace Assay demonstrated great concordance in detecting positive samples under low tumor percentage with a 100% positivity for samples with ≥15% tumor, 97% positivity from 10% to15%, and 92% positivity from 5% to10% tumor percentage (Fig. 3C). Additionally, 5 tests were scored MSI-H at tumor percentages below 1% (Supplemental Table S8). These results indicate reliable MSI detection by the VarTrace assay even at low tumor content.

### Analytical Performance of VarTrace Assay Using Synthetic Samples

The analytical sensitivity and specificity of the VarTrace assay was evaluated using two types of synthetic samples: Seraseq® MSI Reference Panel Mix AF5% and internally generated VarTrace positive controls (5%VAF) and negative controls. The VarTrace assay demonstrated 100% sensitivity and 100% specificity in detecting all 5 markers when tested with both 2 ng input (1 ng per reaction tube) of the Seraseq reference samples and 10 ng of the internal controls (Table 3, Supplemental Table S9 and Fig. S33-34). No false positive results occurred for any markers in the negative control samples in either analytical study.

**Table3.** Analytical Performances of VarTrace using Synthetic Samples.

| Analytical Validation |  |  |
| --- | --- | --- |
| Samples | Seraseq MSI Reference | VarTrace Internal Ctrl |
| Input | 2 ng | 10 ng |
| Specificity | 100% (CI: 83% to 100%) | 100% (CI: 83% to 100%) |
| Sensitivity | 100% (CI: 83% to 100%) | 100% (CI: 83% to 100%) |

| Limit of Blank: 10 ng input |  |  |  |
| --- | --- | --- | --- |
| Samples | WT (+3) 100%VAF | WT (0) | WT (-3) 100%VAF |
| Concordance | 12/12(100%) | 12/12(100%) | 12/12(100%) |

| Limit of Detection: 10 ng input |  |  |
| --- | --- | --- |
| Concordance at Samples | Synthetic Ref Material (-5) | Synthetic Ref Material (-7) |
| 50%VAF | 6/6 (100%) | 6/6 (100%) |
| 10%VAF | 6/6 (100%) | 6/6 (100%) |
| 5%VAF | 4/6 (67%) | 6/6 (100%) |
| 1%VAF | 0/6 (0%) | 4/6 (67%) |

| Limit of Input |  |  |
| --- | --- | --- |
| Concordance at Input | Synthetic Ref Material (-7) 5% VAF | Seraseq MSI Reference 5%VAF |
| 1ng | 6/6 (100%) | 6/6 (100%) |
| 0.5ng | 6/6 (100%) | 6/6 (100%) |
| 0.25ng | 6/6 (100%) | 6/6 (100%) |
| 0.1ng | 5/6(83%) | 6/6 (100%) |

To assess the limit of blank, synthetic wild-type (WT) fragments for 5 markers were tested in the following groups: WT (0), WT (+3) with 3 nt addition, and WT (−3) with 3 nt deletions in the mononucleotide repeat region. Across 12 repeats of each WT group, no WT genotypes demonstrated positive signals for any markers, indicating 100% correct negative scoring within the wild-type allele length range (Table 3, Supplemental Table S10 and Fig. S35-36).

The limit of detection was evaluated using variants with 7 nt deletions in the mononucleotide repeat regions of all 5 markers (MSI-7 variant pool) at VAFs ranging from 1% to 50%. These variants were designed to be near the WT allele length range and difficult to distinguish from WT. The results showed the VarTrace assay could detect and score all experimental repeats containing the −7 variant at 5% VAF as positive (Table 3, Supplemental Table S11 and Fig. S37-40). This is consistent with detection of the BAT-25, NR-24 and MONO-27 variants with 6 nt deletions present at 5% VAF in the Seraseq reference sample from the analytical study.

The limit of input was assessed using the Seraseq Positive Reference (5% VAF) and MSI-7 variant pool (5% VAF), as sensitive samples representing low tumor content with short microsatellite deletions. Input amounts of 1 ng to 0.1 ng per tube of these 2 samples were tested in 6 repeats. The VarTrace assay yielded positive results for all 5 markers in all repeats with as little as 1 ng input (0.5 ng per tube) for both samples at 5% VAF (Table 3, Supplemental Table S12 and Fig. S41-42). Evaluating MSI classification, the VarTrace assay detected MSI-H status with occasional marker drop-out at all repeats of both samples down to 0.5 ng input (0.25 ng per tube).

## Discussion

VarTrace is a qPCR assay that can detect MSI status using 5 markers from Bethesda Guidelines directly from tumor sample DNA without matching normal sample. Our study demonstrated 100% sensitivity and 98% specificity for the VarTrace assay compared to the OncoMate assay across 101 FFPE samples. VarTrace also showed better performance than OncoMate when benchmarked to vendor pathology reports. Beyond comparable accuracy and sample requirement benefit, VarTrace also showed advantages in industrial adaptability over OncoMate. Unlike the two-step pre-amplification and capillary electrophoresis process required for OncoMate, the VarTrace assay platform enables direct qPCR readout from the sample. This simplification of the workflow reduces instrumentation requirements, as shown through comparison of the timelines in Fig. 2E. This enabled VarTrace to be completed in 8 hours, faster than the 10+ hour OncoMate timeframe and better suited for shift work. The higher throughput scalability of qPCR vs capillary electrophoresis also enables VarTrace to process 3-fold more samples per run (Table 2).

Studies of tumor percentage dilution and analytical performance using synthetic samples showed the VarTrace MSI Assay could detect MSI with 97% positivity at tumor content as low as 10% and DNA input as little as 0.5 ng.

However, when markers were considered individually, the sensitivity and specificity of NR-24 was lower at 80% and 90% respectively compared to OncoMate. Discordant NR-24 results showed deletions of 4-7 nucleotide mononucleotide repeats. In an additional limit of detection study using an MSI-5 10% variant pool (Supplemental Table S11), VarTrace also demonstrated 2 false negatives for NR-24 out of 6 repeats. This indicates less sensitive detection of short deletions less than 7 nt by NR-24 in the VarTrace assay. However, NR-24 did show positive amplification without reaching the calling threshold in these discrepant samples. Re-evaluating the NR-24 calling threshold could potentially improve accuracy of this marker.

Next-generation sequencing (NGS) panels can concurrently detect MSI status and alterations in MMR genes from solid tumor samples. FoundationOne CDx (24), as a representative NGS panel, became the first FDA-approved NGS test for pan-cancer biomarker profiling and tumor mutational burden, including multiple microsatellite markers to determine MSI status. It also enables discovery of novel microsatellite markers across cancer types (3, 4, 20, 24, 26, 28), ovarian cancer (24) for instance. While NGS provides additional benefits, its use for first-line MSI testing in colorectal cancer may be prohibitively expensive and has limited clinical utility beyond determination of MSI status.

The VarTrace assay platform has potential for broader applications through validation of additional MSI markers identified by NGS, as well as alternative sample types like cell-free DNA (cfDNA) (25, 34, 35). cfDNA enables minimally invasive MSI testing compared to tissue biopsies, but detecting MSI in cfDNA requires even higher sensitivity due to limited tumor DNA in plasma. VarTrace can better enrich and amplify minimal MSI markers in cfDNA by simply suppressing *GAPDH* amplification to minimize consumption of PCR reagents by this control gene. With further optimization and calibrated calling thresholds, the same platform may be adapted for sensitive and specific MSI detection from cfDNA pending clinical validation. Expanding the validated markers and sample types would extend the utility of the rapid, robust VarTrace assay for MSI analysis across clinical settings.

In conclusion, our findings demonstrate that the VarTrace Assay is a promising new tool for fast, accurate and broadly applicable MSI testing across clinical settings. This assay stands poised to redefine the standard for MSI detection, extending its relevance not only to colorectal cancer but potentially to other applications as well. The implementation of this assay holds the potential to facilitate Lynch syndrome screening on a grand scale, significantly reduce turnaround times for surgical decision, and make possible the assessment of MSI status even in constrained biopsy samples or cfDNA. Further clinical validation and implementation studies will help translate these analytical performance improvements into meaningful patient benefits.

## Data Availability

All data produced in the present study are available upon reasonable request to the authors

## Acknowledgments

The authors thank Dr. Mingjie Dai for valuable suggestions to the manuscript. The authors wish to acknowledge the assistance of Jana Harvey and Jonathan Tran in experiment execution and material organization for this study.

## Ethic Statement

This study was conducted in accordance with the Declaration of Helsinki. The research protocol was approved by the Institutional Review Board at NuProbe USA Inc (IRB number: NP257).

## Preprint Statement

This work has also submitted as preprint at medRxiv.

Link: https://medrxiv.org/cgi/content/short/2023.11.07.23298217v1

DOI: https://doi.org/10.1101/2023.11.07.23298217

